# Short-term outcomes of the digital combined lifestyle intervention CooL-MiGuide: a descriptive case series study

**DOI:** 10.1101/2024.01.17.24301417

**Authors:** Nicole Philippens, Ester Janssen, Stef Kremers, Rik Crutzen

## Abstract

**Introduction:** The objective of this study is to investigate the changes over time in anthropometrics, behaviours and health perceptions after eight months of participation in the Combined Lifestyle Intervention (CLI) CooL-MiGuide, a digital version of the Coaching on Lifestyle (CooL) intervention with the add-on of a digital platform and an app.

**Method:** Longitudinal data were collected between January 2022 and December 2023 from adults, mainly in the South-Western area of the Netherlands, that meet the inclusion criteria for the CLI (i.e. being obese (Body Mass Index (BMI)>30) or being overweight (25<BMI<30) combined with comorbidity; and being sufficiently motivated). We collected a broad set of data at baseline (T0) and after eight months of participation (T1) on anthropometrics (self-reported weight, length, waist circumference), control and support (self-mastery, social support), physical activity (sedentary time on least/most active days, physical active minutes), diet attentiveness (attentiveness to meal composition, amounts of food and consuming), alcohol consumption, smoking, perceived fitness (perceived health, fitness when waking, fitness during daytime, impact daily stress), sleep and stress. Changes over time were analyzed using paired t-tests comparing T1 to baseline. Effect sizes were calculated using Cohen’s d.

**Results:** The results show positive changes after eight months in anthropometrics, sedentary time, diet attentiveness, sleep, stress, fitness, and perceived health of the participants. Largest effect sizes were found for weight (Cohen’s d=0.62, average weight loss of 3.82kg), perceived health (Cohen’s d=0.58) and perceived fitness of the participants when waking and during daytime (Cohen’s d=0.41 and 0.52 respectively).

**Conclusion:** CooL-MiGuide participants show relevant changes in the essential constructs perceived health and weight. The digital version of CooL can be a valuable alternative for people who prefer participation from a location of their choice.

## Introduction

In 2022, 50.2% of Dutch people aged 18 years and older were overweight and 15.1% were obese [1]. Obesity is considered a disease according to the World Health Organisation and the Dutch Health council [2, 3].

A Combined Lifestyle Intervention (CLI) is a health intervention for people with overweight or obesity, coaching participants towards a healthier lifestyle. A CLI exists of group and individual sessions over a course of 24 months, covering the topics of healthy diet, physical activity, and behavioural change. As of January 2019, Combined Lifestyle Interventions (CLIs) are part of basic health insurance in the Netherlands.

The Coaching on Lifestyle intervention (CooL) is one of six CLIs that are approved and accredited by the Dutch Institute for Public Health and Environment (in Dutch: Rijksinstituut voor Volksgezondheid en Milieu) for being effective in facilitating weight reduction. The CooL-intervention is usually offered in physical mode (i.e., by means of face-to-face contact). The COVID-19 pandemic, entering the Netherlands in February 2020, caused a shift in CooL-execution modus from physical towards digital, providing additional challenges for both coaches and participants [4]. CooL-MiGuide is a digital version of CooL that was initiated during the COVID-19 pandemic and was continued afterwards as requested by CooL-referrers, care groups and CooL-participants. From the perspective of participants, the digital version of CooL is easy to use, less time consuming and appealing to a certain segment of the public (e.g., for participants providing informal care or childcare, for less mobile participants or participants with irregular working hours or locations) [5].

In short, the inclusion criteria for CLIs in the Netherlands are being obese or overweight with comorbidity, being 18 years or older and being sufficiently motivated to complete the two-year intervention. A detailed description of the CLI-inclusion criteria is available elsewhere [6]. For CooL-MiGuide, additional inclusion criteria apply, e.g., participants must be digitally skilled, master the Dutch language and be in possession of a smartphone and laptop, tablet or computer. Participants that apply for CooL-MiGuide but do not meet these additional inclusion criteria, as noticed by both participant and staff during the introductory phone call, are referred back to their general practitioner for participation in CooL (with face-to-face sessions) or other suitable care.

The objective of this study is to investigate the effect of a digital version of CooL on the changes over time in anthropometrics, behaviours and health perceptions after eight months of participation.

## Method

### CooL

CooL aims for higher perceived quality of life, healthier eating habits, more physical activity, less sedentary behaviour, attention for high quality sleep and relaxation, resulting in positive changes in physical outcomes such as weight, BMI and waist circumference. The monitoring of CooL covers all areas with perceived health and weight as the primary outcome measures.

CooL includes an intake (1 hour), a behavioural change phase of eight months (phase 1) and a follow-up phase of sixteen months (phase 2). The intervention consists of a combination of individual sessions and 16 group sessions (1,5 hours each) all led by one and the same coach. Phase 1 and phase 2 both include eight group sessions with a higher density of sessions in phase 1 compared to phase 2. The CooL-coaches are trained and licensed professionals who coach participants to take responsibility for their personal lifestyle changes by addressing motivation, personal objectives and behavioural change. Participants are stimulated and supported towards more self-steering and self-management by identifying, planning, and putting personal health related behaviour into action. The main objective of CooL is to coach and activate participants to a sustained healthier lifestyle in line with their individual needs and goals. A detailed description of the CooL-intervention and data collection method is available elsewhere [6].

### CooL-MiGuide: a digital version of CooL

CooL-MiGuide is a digital version of the CooL-intervention pursuing the same aims as CooL. In CooL-MiGuide, all sessions are offered in digital modus (videoconferencing) and participants can make use of a supporting MiGuide-app and MiGuide-portal offering synchronous and asynchronous communication options with the CooL-coach and other group members, facilitating and stimulating mutual contact. In addition, the MiGuide-portal provides additional sources of information that are easily and at any time accessible, whereas the MiGuide-app provides participant specific information and challenges.

The working elements of CooL-MiGuide are for the major part similar to CooL. The similarities and differences in working elements between both versions have been thoroughly investigated and are elaborated in detail [7]. The CooL-MiGuide coach pays specific attention to working elements that may differ from the physical mode of the intervention, mainly related to the interaction between coach and participant(s) and among participants, i.e., experiential learning, mutual connection, and cooperation. Additionally, the regional connection of CooL-MiGuide and local health and welfare organizations, as well as the connection with local sport facilities requires extra attention. On the other hand, CooL-MiGuide offers additional working elements compared to CooL, by offering additional cues to action (via prompts, invitations to challenges, the use of behavioral diaries and self-measurements in the MiGuide-app and MiGuide-portal), more (digital) contact moments and more flexibility through participation from a location of choice (e.g. home, workplace).

### Study design and population

As CooL is part of regular health care, a study design with a control group receiving no treatment would be unethical. CooL-MiGuide was initiated during the COVID-19 pandemic, during which a temporary extension in the CLI-regulations on health insurance, made it possible to offer CooL digitally instead of via face-to-face contact only [8]. Both the digital and face-to-face version of CooL were maintained in the aftermath of COVID-19, which makes it possible to look simultaneously at outcomes of both delivery modes of CooL.

The participants, all Dutch-speaking adults living in the Netherlands, were included from January 2022 until December 2023, at different locations mainly in the (South-)Western part of the Netherlands. All but one participant met the inclusion criteria for participating in a CLI. In one case (n=1, 0.78 %), BMI and waist circumference at baseline were below the inclusion threshold, potentially due to lifestyle changes in the time between participant’s application and the start of CooL. Since the waist circumference of this participant was above the threshold at the moment of referral, this case was included.

### Power

The average effect size on the change in perceived health and weight in prior CooL-related research (0.52 and 0.56 respectively [4, 9] was used as input for the power analysis. Based on d=0.52 for a difference between two dependent means, a two-tailed t-test with an alpha of 0.05, and a minimal power of 0.80, the required sample size is 32 participants.

### Data collection

For CooL-MiGuide-participants we used the same questionnaire as for CooL-participants. The outcome measures we collected can be categorized into anthropometrics (i.e., weight/BMI and waist circumference), control and support (i.e., self-mastery and social support), physical activity (i.e., sedentary time on least/most active days and active minutes), diet attentiveness (attentiveness to meal composition, awareness to amounts of food and attentiveness to consuming), alcohol use and smoking, perceived fitness (i.e., perceived health, perceived fitness when waking, perceived fitness during daytime and impact of stress on daily functioning), sleep and stress. A more detailed description of both questionnaire and anthropometrics is available elsewhere [6].

Data were collected at three time points during CooL-MiGuide: at the beginning of the intervention, during the intake (T0); after 8 months, at completion of phase 1 of the intervention (T1); and after 24 months, at completion of the intervention (T2). Data from T2 were not yet available at the time of the analysis and are not presented in this article.

Due to the online nature of CooL-MiGuide, anthropometrics (i.e. body weight and waist circumference) were measured by the participant instead of the CooL-coach. All participants received instructions how to perform both measurements accurately. The use of the MiGuide-application stimulates participants to perform additional anthropometric measurements in between sessions in order to follow-up on their progress.

### Analyses

Variables were recoded in line with the approach used in the validated questionnaires and to facilitate interpretation of the outcomes (i.e., direction of effect sizes). In addition, we performed an exploratory factor analysis and calculated McDonald’s omega to assess the internal structure of items regarding the constructs using R software [10]. These analyses justified summarizing the constructs by item score means. For all items and constructs we ran descriptive analyses (e.g., means, standard deviations). Changes in outcome measures over time were analysed using paired t-tests (T1 versus T0). Effect sizes were calculated using Cohen’s d and the outcomes were interpreted in accordance with Lipsey’s guidelines for each pair of outcomes, i.e., an effect size smaller than or equal to 0.32 is considered small, an effect size between 0.33 and 0.55 is considered medium and an effect size of 0.56 or above is considered large [11]. To improve comprehensibility effect sizes are represented such that positive values represent change in the desired direction whereas negative values represent change in the undesired direction.

To be considered successful, the target for the CLI (including CooL) is a 5% weight loss after the two years intervention, as set by the Dutch Healthcare Institute (Dutch: Zorginstituut) based on the guidelines set up by the Dutch Institute for Quality in Health Care (Dutch: Centraal Begeleidingsorgaan) as well as their English counterpart (National Institute for Health and Care Excellence) [12]. The data in this study covers the first phase of CooL-MiGuide only (8 months), still leaving 16 months to further extend weight loss. We categorized the outcomes on weight change: 5% weight loss or more, between 0 and 5% weight loss, weight stabilization or weight gain.

### Ethics

This study was submitted to and approved by the Research Ethics Committee of the Faculty of Health, Medicine and Life Sciences of Maastricht University (FHML-REC/2019/073). All participants gave their informed consent for their anonymised personal data to be used for research purposes.

### Funding

No funding was provided for this research.

## Results

In the result section all outcomes are displayed starting with the demographics of the participants (Table 1) and followed by outcome measurements (Table 2), including mean values and standard deviations as well as confidence intervals on changes in outcomes and effect sizes.

**Table 1.**
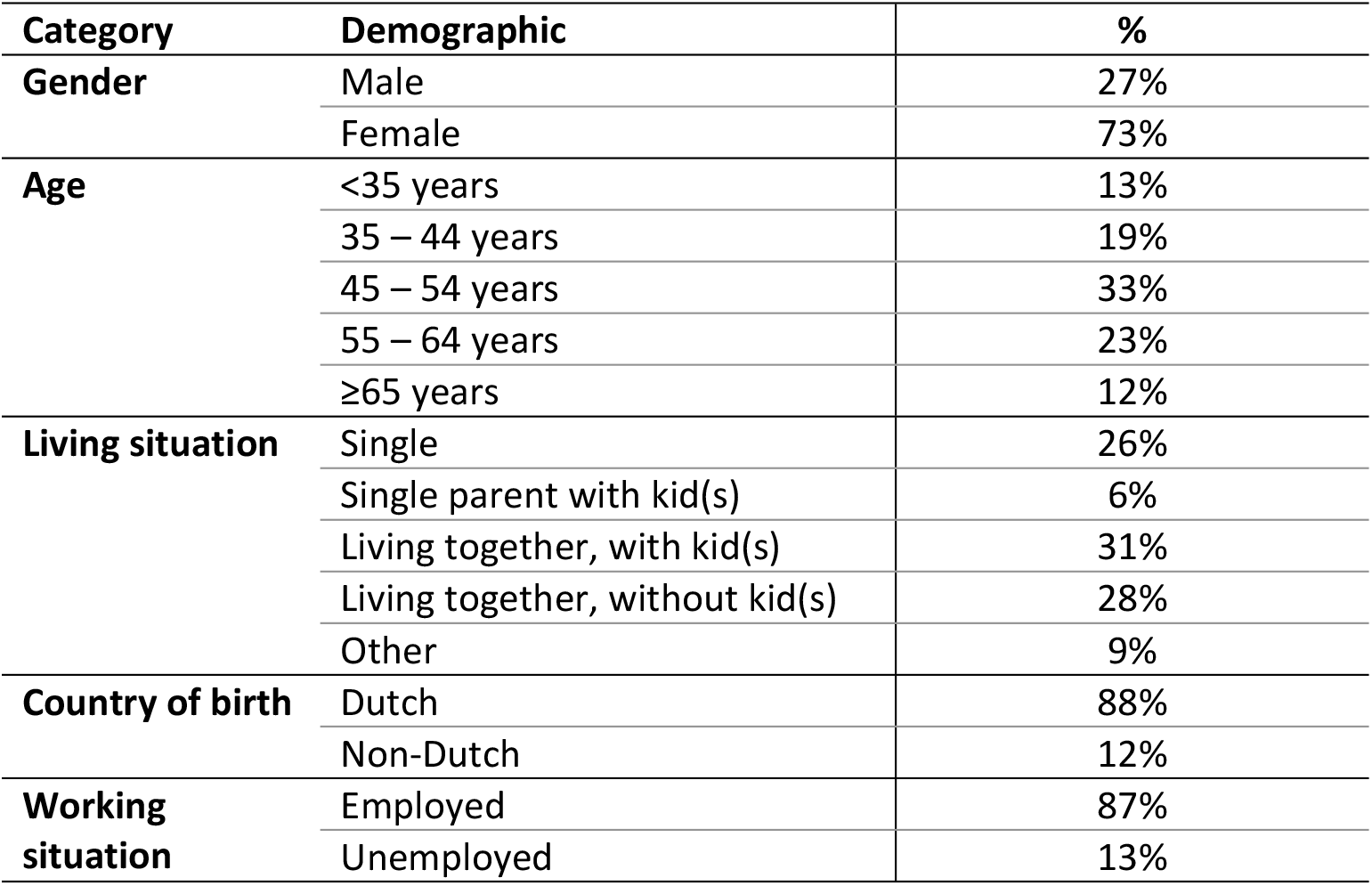
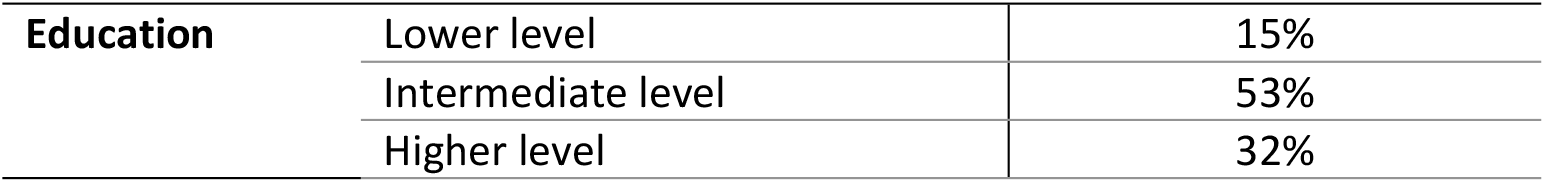
Demographics of CooL-MiGuide participants (N= 129).

**Table 2.** Results on all CooL-MiGuide outcome measurements (N=129).

| Category | Construct / factor | T0 M (sd) | T1 M (sd) | $\Delta T0-T1$ [95% CI] | d** |
| --- | --- | --- | --- | --- | --- |
| Anthropo-metrics | Weight | 108.66 (23.47) | 104.94 (23.03) | -3.82 [-4.91; -2.72]* | 0.62 |
|  | BMI | 36.62 (6.96) | 35.33 (6.78) | -1.32 [-1.69; -0.95]* | 0.63 |
|  | Waist circumference | 115.53 (15.10) | 113.32 (15.77) | -2.37 [-3.91; -0.83]* | 0.28 |
| Control & support | Self-mastery | 3.30 (0.70) | 3.35 (0.63) | 0.04 [-0.07; 0.15] | -0.07 |
|  | Social support | 3.61 (0.93) | 3.72 (0.79) | 0.10 [-0.04; 0.23] | 0.12 |
| Physical activity | Sedentary time (least active) | 8.67 (3.34) | 8.01 (2.90) | -0.67 [-1.18; -0.15]* | 0.23 |
|  | Sedentary time (most active) | 5.30 (2.94) | 4.55 (2.58) | -0.76 [-1.19; -0.33]* | 0.31 |
|  | Physical active minutes | 82.53 (101.82) | 90.22 (112.32) | 7.69 [-11.00; 26.38] | 0.07 |
| Diet attentiveness, alcohol and smoking | Attentiveness for meal composition | 3.20 (0.87) | 3.52 (0.65) | 0.32 [0.17; 0.48]* | 0.36 |
|  | Awareness for amounts of food | 3.15 (0.87) | 3.52 (0.71) | 0.37 [0.20; 0.55]* | 0.37 |
|  | Attentiveness to consuming | 2.98 (0.93) | 3.26 (0.79) | 0.28 [0.13; 0.42]* | 0.34 |
|  | Alcohol consumption | 1.37 (2.20) | 1.37 (2.16) | 0.00 [-0.34; 0.34] | 0.00 |
|  | Smoking*** | 0.63 (3.02) | 0.29 (2.00) | -0.35 [-0.73; 0.03] | 0.16 |
| Perceived fitness | Perceived health | 8.64 (2.06) | 9.74 (1.67) | 1.10 [0.77; 1.43]* | 0.58 |
|  | Fitness (when waking) | 2.13 (0.80) | 2.46 (0.79) | 0.33 [0.19; 0.47]* | 0.41 |
|  | Fitness (during daytime) | 2.21 (0.78) | 2.60 (0.78) | 0.38 [0.26; 0.51]* | 0.52 |
|  | Impact stress (daily) | 2.02 (0.91) | 2.22 (0.98) | 0.20 [0.05; 0.36]* | 0.23 |
| Sleep | Sleep (summary) | 12.40 (3.68) | 11.50 (3.34) | -0.86 [-1.40; -0.31]* | 0.27 |
| Stress | Stress (summary) | 19.00 (5.82) | 17.98 (5.60) | -0.96 [-1.74; -0.19]* | 0.22 |

\*p<0.05
\*\*Positive effect size representing an effect in desired direction, negative effect size representing an effect in undesired direction
\*\*\* The dataset contains eight smokers at T0 and six remaining smokers at T1.

### Demographics

A total of 129 adults started with CooL-MiGuide between Feb-2022 and May-2023, completed phase 1 of CooL-MiGuide before 2024 and returned the T0 and T1 questionnaires. The amount of missing data differed per item per time of measurement (range 0.8-8.5%).

Of all participants a total of 27% were male and 73% female. Most participants (88%) had a Dutch background. A quarter of the participants was living alone and 65% was living with children and/or partner, 87% were employed and two third of the participants had a lower or intermediate level of education (Table 1).

### Anthropometrics

Weight, BMI and waist circumference all showed a decrease after eight months (T1) compared to baseline (T0). The BMI of the participants was on average 36.6 at baseline and decreased with 1.3 BMI-points at T1. The average weight loss was 3.8 kg at T1, corresponding to a 3.5% average weight loss per participant after eight months. In total 80% of the participants lost weight and 28% of the participants lost over 5% body weight at T1. The average waist circumference of the participants decreased from 115.5 cm at T0 to 113.3 cm at T1. The change in waist circumference demonstrated a small effect size (Cohen’s d=0.28) at T1, whereas weight and BMI showed a large effect size at T1 (0.62 and 0.63 respectively).

### Control and support

Self-mastery and social support showed no changes at T1 compared to baseline.

### Physical activity

Sedentary time decreased at T1 both for least and most active days of the week: participants sat on average 40 minutes less on least active and 46 minutes less on most active days, compared to baseline. The effect size on sedentary time was small (between 0.23 and 0.31). The average daily active minutes showed no changes at T1 compared to baseline.

### Diet attentiveness, alcohol and smoking

Over time the participants showed an increase in attentiveness for meal composition, awareness for the amounts of food selected and attentiveness when consuming food. The effect size on all three items was medium-sized (0.36, 0.37 and 0.34 respectively) when comparing baseline to T1. 74% of the participants indicated a healthier or much healthier eating pattern, 25% indicated no change and 1% ate less healthy at T1 compared to baseline. Alcohol consumption and smoking showed no change at T1 compared to baseline, though the number of smokers decreased from eight to six between baseline and T1.

### Perceived fitness

The indicators for perceived fitness, i.e. perceived health, feeling fit when waking up, feeling fit during the day and the impact of stress on daily functioning, all showed an effect in the desired direction. The effect size was small for the impact of stress on daily functioning (0.23), medium for feeling fit when waking up and feeling fit during the day (0.41 and 0.52 respectively) and large for perceived health (0.58).

### Sleep and stress

The constructs sleep and stress both showed a decrease at T1 compared to baseline with a small effect (0.27 and 0.22 respectively) in the desired direction.

## Discussion

This is the first study on CooL fully offered in digital mode in combination with the MiGuide-app and MiGuide-platform. The results of CooL-MiGuide show positive changes in self-reported anthropometrics, behaviours (sedentary time, diet attentiveness) and perceptions (sleep, stress, fitness, and health) of the participants after eight months. Largest effect sizes were found on weight (0.62)/BMI (0.63), and perceived health (0.58). Medium effect sizes were found on diet attentiveness (between 0.34-0.37) and perceived fitness (between 0.41-0.52).

The set-up, approach and content of CooL is adopted in CooL-MiGuide, ensuring a similar intervention, enabling comparison on delivery mode. When comparing CooL-MiGuide with earlier research on CooL, a notably similar pattern in outcomes is visible: small to medium effect sizes in behaviours and awareness, and relatively large effect sizes in weight and perceived health [4, 9].

The effect size on waist circumference (0.28) does not keep pace with the effect size on weight (0.62) and remains smaller compared to earlier research on CooL [4, 9]. Preferably, waist circumference measurements are done by professionals, as is the case for CooL, since these measurements require a strict protocol, training and repeated measurements [13]. Measuring waist circumference accurately on an occasional basis by non-professionals is challenging. For CooL-MiGuide these measurements were carried out by participants themselves, potentially creating a larger margin of error. Still, a change can be detected, as demonstrated by the outcomes of CooL-MiGuide, but is simply more difficult to capture (i.e., reduced sensitivity). The weight measurements, done by the participants themselves on ordinary home equipment, generate lower reliability as well, but potential deviations in the equipment are unlikely within the timeframe of eight months.

CooL-MiGuide-participants reported to have reduced their sedentary time and felt more fit when waking and during the day, whereas on physical activity (PA) no effect was visible. The results, a decrease in sedentary time and limited or no effect on PA, the latter in combination with large confidence intervals, are in line with earlier research on CooL [4]. Requesting input on minimum bouts of PA of 10 minutes, as well as the general perception of PA (i.e., being intensive) might raise barriers for registration. However, the decrease in sedentary time is already promising, as sedentary time is likely to be displaced by light intensive activity, generating a positive impact on health [14]. A finding that seems to be supported in this study by the improved outcomes on all constructs of perceived fitness for CooL-MiGuide-participants.

The CooL-MiGuide-sample, compared to previous studies on CooL, shows a similar gender distribution, but contains more participants with a non-Dutch background, more participants who are single, with a job and participants with an intermediate-level of education. Participants with a lower level of education are less represented. A process evaluation on the CooL-MiGuide-implementation in June 2023 showed that as much as 92% of the applicants for CooL-MiGuide effectively started with the intervention [5]. Reasons for not starting were mostly independent of the delivery mode of CooL-MiGuide (insufficient knowledge of Dutch language, dislike for group sessions, no motivation for intensive program) and partly related to the digital version (disliking digital interventions). However, a larger number of participants is needed to shed light on the sociological differences between participants attracted by the different delivery modes of CooL.

The complementary offering of CooL-MiGuide might positively impact the motivation of the participants. Providing participants with a choice between a physical or digital program mode, contributes to the perception of self-control and autonomy and assures a better fit with the participants needs and wishes thereby preventing dropout and enhancing motivation to participate.

### Strengths and limitations

This study has several strengths. Firstly, a broad scope of data is collected. Secondly, we were able to investigate a topical and innovative adaptation of an existing evidence-based intervention which enables further testing, development, implementation and scale-up.

There are also some limitations in this study. To begin, CooL-MiGuide does not yet offer nationwide coverage though quite some regions are already in scope. Secondly, due to the digital mode of delivery, the anthropometrics are collected by self-measurement thereby generating a larger margin of error than measurements done by professionals.

### Recommendations for future research

We recommend future research on CooL-MiGuide to address the following topics:

- Study on the long-term effects of the CooL-MiGuide intervention (i.e., including 24 months measurements).
- Study on the characteristics of the participants that are appealed by the digital version of CooL and participants that are not, as well as on the characteristics of participants that drop-out of CooL-MiGuide, in order to maximize digital inclusion when scaling up the intervention.
- Study on the overall dropout rates of CooL when offering the choice between physical and digital mode of CooL.
- Study on the social cohesion within a CooL-MiGuide group of participants compared to similar interventions offering face-to-face contact moments and its impact on the outcomes of the intervention.

## Conclusion

CooL-MiGuide-participants show relevant short-term changes in the essential constructs of perceived health and weight. The digital version of CooL can be a valuable alternative to physical combined lifestyle interventions for people who prefer participation from a location of their choice.

## Data Availability

The datasets generated and/or analysed during the current study are not publicly available because the informed consent statement to using data at the individual level was limited to the authors of this article and are only available from the corresponding author on reasonable request.

https://osf.io/fjctx/?view_only=292e8cb696b842b9804394c81e7703e4

## Abbreviations

BMI: Body Mass Index
CLI: Combined Lifestyle Intervention
CooL: Coaching op Leefstijl
PA: physical activity

## Acknowledgements

The authors like to thank the MiGuide Organization, CooL-MiGuide-coaches and the CooL-MiGuideparticipants for their efforts to make this research possible.

## Notes

### Competing Interest Statement

Not applicable for S.K. and R.C.. Both main authors (E.J. and N.P.) are co-owner of the CooL-intervention.

### Funding Statement

The author(s) received no specific funding for this work.

